# Clinicopathological Factors Associated with Gastric Signet Ring Cell Carcinoma in *CDH1* Pathogenic Variant Carriers: Report from the GASTRIC Consortium

**DOI:** 10.64898/2026.03.27.26349321

**Authors:** Ophir Gilad, Christine M. Drogan, Emma Keel, Guimin Gao, Carol Swallow, Anand Govindarajan, Savtaj Brar, Melissa Heller, Taylor Apostolico, Michelle F. Jacobs, Kebire Gofar, Beth Dudley, Eve Karloski, Conner Lombardi, Michelle Springer, Souvik Saha, Devin Cox, Benjamin A. Lerner, George Hanna, Yana Chertock, Afshin Khan, Sarah Ertan, Kimberly Hilfrank, Sheila D. Rustgi, Aparajita Singh, Michael J. Hall, Xavier Llor, Ajay Bansal, Swati G. Patel, Randall E. Brand, Maegan E. Roberts, Peter P. Stanich, Elena Stoffel, Bryson W. Katona, Melyssa Aronson, Sonia S. Kupfer

## Abstract

**Background:** Gastric cancer surveillance in *CDH1* pathogenic variant carriers is challenging, as predictors of localized (stage T1a) and advanced (stage >T1a) signet ring cell carcinoma (SRCC) are not well defined. We established the **G**roup of investig**A**tors **ST**riving toward **R**esearch **I**n ***C****DH1* (**GASTRIC**) consortium to identify clinicopathological factors associated with localized and advanced SRCC.

**Methods:** A retrospective observational study (1998–2025) of *CDH1* carriers across twelve academic centers was performed. Clinical, endoscopic, and pathological data were compared between carriers with and without SRCC on endoscopy, and between those with advanced versus localized or no cancer on gastrectomy specimens.

**Results:** Overall, 390 *CDH1* carriers from 235 families were included. Presence of SRCCs on endoscopy was significantly associated with thickened folds, nodularity, masses, and intestinal metaplasia, while gastritis was negatively associated. Of 196 carriers (52.4%) undergoing gastrectomy, 11 (5.6%) had advanced cancers, 10(90.9%) of which showed endoscopic abnormalities. Identification of SRCC on baseline endoscopy was the most sensitive feature for advanced disease (0.81) but had moderate specificity (0.74), whereas masses and thickened folds were highly specific (0.99 and 0.96, respectively) but less sensitive. Negative predictive values were high (0.94-1.0), while positive predictive values were modest (0.13-0.66). On multivariate analysis, masses and SRCC foci on baseline endoscopy were independent predictors of advanced disease.

**Conclusion:** Among *CDH1* carriers, absence of endoscopic findings was reassuring, whereas significance of detected endoscopic and pathological abnormalities was less certain. Advanced cancer occurred in a small number of carriers, with endoscopic abnormalities in nearly all cases. Endoscopic surveillance might be an alternative to surgery in carriers without worrisome mucosal findings.

**What You Need to Know:** *Background:* Predictors of advanced stage disease in *CDH1* carriers, which could help in risk stratification and management of these individuals, are not well defined. Identifying associated features could improve risk stratification and management.

*Findings:* Endoscopic and histologic features associated with advanced disease showed variable sensitivity and specificity. Absence of abnormalities had high negative predictive value, whereas presence of findings had limited ability to reliably predict advanced disease.

*Implications for Patient Care:* Absence of worrisome findings is reassuring and may support continued surveillance. Detected abnormalities warrant close evaluation and discussion of surgery, though their presence alsone does not reliably indicate advanced disease.

## Background

*CDH1-*associated syndromes increase lifetime risk of diffuse gastric cancer and/or lobular breast cancer. Historically, lifetime risks of diffuse gastric cancer were estimated at 40-70% depending on family history^1–3^, though a more recent study reported lifetime risk as low as 7-10%^4^. The hallmark of the condition is gastric signet ring cell carcinoma (SRCC) that, in the majority of *CDH1* carriers, is localized to the mucosa (stage T1a) and can remain indolent throughout their lifetime^5,6^. Identification of localized T1a lesions can be challenging endoscopically due to absence of or only subtle mucosal abnormalities. Recent single center studies suggest endoscopic surveillance is safe and effective in *CDH1* carriers, though larger studies and longer follow-up are needed^7,8^.

A subset of *CDH1* carriers progress to more advanced (stage >T1a) disease with poorer outcomes. While prophylactic total gastrectomy (TG) has been the gold standard for reduction of advanced diffuse gastric cancer risk, not all *CDH1* carriers require surgery due to variable penetrance. However, predictors of advanced stage disease are not well defined. Identification of clinical, endoscopic and/or pathological factors associated with advanced versus localized or no SRCC would be an important advance to inform clinicians and patients in their shared decision-making about optimal management.

To address these gaps in the field, the **G**roup of investig**A**tors **ST**riving toward **R**esearch **I**n **C**DH1 (**GASTRIC**) consortium was established and collected harmonized retrospective data on *CDH1* pathogenic variant carriers followed at 12 academic centers in the US and Canada between 1998 and 2025. This consortium is among the largest cohorts of *CDH1* pathogenic variant carriers. In this first report from the GASTRIC consortium, the aims were to characterize clinical, endoscopic and histologic factors associated with presence of SRCC foci on endoscopic biopsies, as well as predictors of advanced stage versus localized stage or no disease on TG specimens.

## Methods

### Clinical data

Individuals with a pathogenic or likely pathogenic variant in *CDH1* were identified from 11 academic centers in the United States and 1 center in Canada. The study was approved by the institutional review boards of all centers. Retrospective data were collected from the electronic health record and included age, sex, race and ethnicity, family history of gastric and breast cancers, proton pump inhibitors (PPI) use, smoking status (never, past or current smoker) and alcohol consumption (none, occasional or heavy). Classification of *CDH1* variant was collected from genetic test reports, and age of diagnosis was defined as age at identification of the *CDH1* variant. When individuals from different sites shared the same *CDH1* variant, their pedigrees were reviewed to avoid duplicate entries. Clinical criteria were assessed according to both the 2020 International Gastric Cancer Linkage Consortium (IGCLC guidelines)^1^ and the 2025 National Comprehensive Cancer Network (NCCN) guidelines^9^ (**Supplementary Table 1**). Carriers with metastatic gastric cancer at presentation were excluded from further analysis.

### Endoscopic data

Procedure and pathology reports from individuals who underwent one or more esophagogastroduodenoscopy (EGD) were reviewed. All endoscopic and histologic features were prespecified during meetings among the principal investigators of the GASTRIC consortium prior to initiation of data collection. Endoscopies were defined as “baseline” if they were the first recorded EGD in the medical record and as “surveillance” for all subsequent EGDs. Endoscopic features were collected from procedure reports and included pre-defined descriptors of thickened mucosal folds, pale mucosal spots, erythema, friability, nodularity, erosions, ulcers, polyps or masses. The number and location of random and targeted biopsies were recorded. The proximal stomach was defined as cardia, fundus and body, while the distal stomach was defined as the transition zone and antrum. Histologic features were collected from pathology reports and included descriptors of gastric intestinal metaplasia (GIM), gastritis, fundic gland polyps, as well as the presence and number of SRCC foci.

### Surveillance data

Carriers undergoing surveillance (i.e. > 1 EGD) were compared to carriers undergoing only a single baseline EGD by demographics, endoscopic findings and surgical data. Oncologic data were not compared as none of the surveillance cohort had advanced cancer.

### Surgical data

In carriers who underwent TG, pathology reports from each site were reviewed for presence of SRCC foci as well as number of foci, size of foci (in mm) and depth of invasion. Carriers with no SRCC or only T1a foci (defined as SRCCs restricted to lamina propria) were considered the localized group, while those with stage>T1a (defined as tumor infiltrating beneath mucosa) were considered advanced.

### Oncological data

Records from carriers diagnosed with advanced stage cancer were reviewed in depth for the presence of any abnormal signs and/or symptoms prior to diagnosis, including abdominal pain, melena, hematemesis, early satiety, anemia or any other clinically significant abnormality determined by the reviewing provider. Chemotherapy and radiotherapy were recorded. Cancer recurrence and death were documented; otherwise, the most recent follow-up showing no evidence of disease (NED) was noted.

### Statistical analyses

Continuous variables are presented as median and interquartile range (IQR) and categorical variables as proportions. Generalized estimating equation (GEE) method^10^ was used to analyze repeated measures from multiple endoscopies and assess associations between clinical, endoscopic or histologic variables and identification of SRCC foci on EGD biopsies. Mann–Whitney test was used to evaluate associations between continuous variables and the presence of advanced disease on TG. Chi-square or Fisher’s exact tests were used for categorical variables. Multivariable analysis of these factors was conducted using logistic regression. Sensitivity, specificity, positive and negative predictive values were calculated to assess the performance of endoscopic features in predicting advanced cancer. Spearman rho was used to evaluate the correlation between number of SRCC foci on EGDs and TG specimens. p value < 0.05 was considered statistically significant for all analyses. We did not adjust for multiple comparisons since *CDH1*-associated syndromes are rare, and advanced cancers in this population is even less common; formal correction would substantially reduce power and increase the risk of type II error. All endoscopic and histologic features were prespecified based on biological plausibility and prior literature, and analyses were hypothesis driven. We therefore emphasize effect sizes, confidence intervals, and clinical relevance rather than adjusted p-value thresholds^11^. SAS version 9.4 was used for GEE analysis, and SPSS software was used for all other analyses (IBM version 25, 2017).

## Results

### GASTRIC consortium patient characteristic

A total of 390 *CDH1* pathogenic/likely pathogenic variant carriers from 235 families were identified (**Table 1**). Median age at *CDH1* diagnosis was 44.6 years (IQR 31.2-56.4), and 143 (36.6%) carriers were male. Median year of diagnosis was 2018 (IQR 2016-2021, range 1998-2025). Most carriers, 329 (84.3%), were non-Hispanic White, while 24 (6.1%) were Hispanic, 17 (4.3%) were non-Hispanic Asian, and 10 (2.5%) were non-Hispanic Black. Full pedigree data was available in 379 individuals, of which 166 (43.9%) met 2020 IGCLC testing criteria, and 310 (82.1%) met the 2025 NCCN testing criteria. The most common variant types were frameshift (37.4% of families), nonsense (16.1%) and intronic (14.9%). Seventeen carriers (4.3%) with metastatic gastric cancer at presentation were excluded from further analyses. A study flowchart is shown in **Figure 1**.

**Figure 1.**
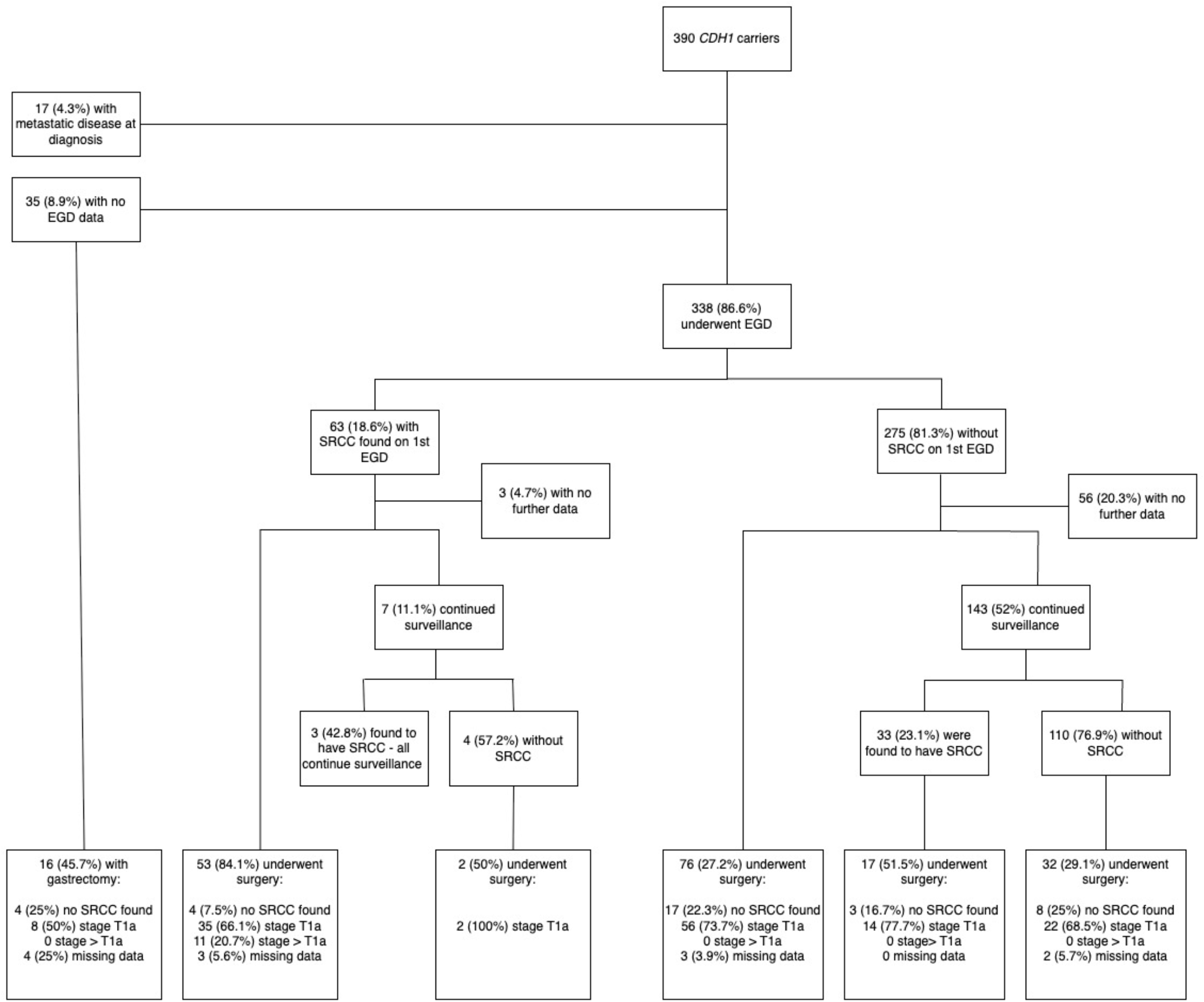
Study flowchart

**Table 1.** Baseline characteristics of *CDH1* carriers in the GASTRIC consortium cohort (n=390)

|  |  |
| --- | --- |
| Age at *CDH1* diagnosis, median, IQR | 44.6 (31.2-56.4) |
| Male sex (%) | 143 (36.6) |
| Race (%) |  |
| Non- Hispanic White | 329 (84.3) |
| Non- Hispanic Asian | 17 (4.3) |
| Non-Hispanic Black | 10 (2.5) |
| Other/missing | 11 (2.8) |
| Ethnicity – Hispanic (%) | 24 (6.1) |
| Smoking ever (%) | 103 (26.4) |
| Pack years, median, IQR | 7.5 (3.5-20) |
| Heavy alcohol consumption (%) | 17 (4.3) |
| PPI use (%) | 104 (26.6) |
| IGCLC criteria met (%) | 166/379 (43.8) |
| NCCN criteria met (%) | 311/379 (82.1) |
| *CDH1* variant classification (% of 235 families) |  |
| Frameshift | 88 (37.4) |
| Nonsense | 38 (16.1) |
| Intronic | 35 (14.9) |
| Cryptic splice site | 28 (12.6) |
| Large deletion/duplication | 27 (11.9) |
| Missense | 2 (0.8) |
| Initiation codon | 1 (0.4) |
| Missing documentation | 16 (6.8) |
| Personal history of breast cancer (% of females) | 85 (34.1) |
| Number of carriers with metastatic disease at diagnosis (%) | 17 (4.3) |
| Number of endoscopies, overall* | 735 |
| Number of endoscopies per carrier, mean ±SD* | 2.1 ±1.8 |
| Number of carriers undergoing surveillance endoscopies ($\geq 2$ EGDs)* | 152 (40.6) |
| Number of carriers undergoing TG (%)* |  |
| Overall | 196 (52.5) |
| Localized (T1a or no SRCC foci) | 173/196 (88.2) |
| Advanced (>T1a) | 11/196 (5.6) |
| Missing data on depth of invasion | 12/196 (6.1) |
| Number of cancer-related deaths* (%) | 4 (1.1) |

IGCLC – International Gastric Cancer Linkage Consortium; IQR – interquartile range; NCCN – National Comprehensive Cancer Network; SD – standard deviation; SRCC – signet ring cell carcinoma; TG – total gastrectomy
\*Excluding carriers with metastatic disease at diagnosis, n=373

### Endoscopic data

Overall, 338 (90.6%) carriers underwent at least one EGD, with a total of 735 procedures ranging between 1-11 EGDs and with a median of 1 (IQR 1-3) procedures per carrier. Ninety-five carriers (28.1%) were found to have SRCC on their EGDs, of which 57 (60%) were identified on the baseline EGD only, 34 (35.7%) were identified on a surveillance procedure, and 4 (4.2%) had SRCC foci identified on both baseline and surveillance EGDs. Eight (2.3%) carriers had multiple EGDs with SRCCs. Of carriers with SRCC found on an EGD, 24 (25.2%) continued surveillance (median of 4 EGDs). Of these, 2 (8%) eventually underwent prophylactic TG which did not identify advanced cancer, while 22 carriers continued with endoscopic surveillance with no signs of overt cancer.

SRCC foci were found in 156/23,046 (0.6%) of random biopsies, and 21/1,132 (1.8%) of targeted biopsies. Most SRCC foci were identified in the proximal stomach including 29 foci in the cardia, 45 in the fundus and 34 in the gastric body, while 10 in the transition zone and 19 in the antrum (sites of the rest were not documented).

After excluding data from EGDs in carriers with metastatic gastric cancer (n=14) and procedures without biopsies (n=10), data from a total of 711 EGDs were evaluated. **Table 2** compares EGD findings between exams with SRCC (115, 16.1%) versus those without SRCC (596, 83.8%) using GEE model. Thickened mucosal folds, pale mucosal spots, erythema, nodularity and gastric masses were significantly associated with presence of SRCCs. Histologically, gastric intestinal metaplasia (GIM) was identified more commonly in carriers with SRCC, whereas the presence of gastritis was associated with a lower likelihood of SRCC detection. Notable, H. pylori was not identified in any procedures with SRCCs. No clinical parameter including age, sex, significant smoking or alcohol history, PPI use, nor meeting any testing criteria were associated with SRCC. On multivariate analysis, the presence of thickened mucosal folds, nodular mucosa, masses or GIM and higher number of biopsies taken were significantly associated with identification of SRCCs, while gastritis was inversely associated with SRCC.

**Table 2.** Comparison of clinical, endoscopic and histologic factors in *CDH1* carriers with vs. without SRCC foci.

|  | SRCC<br>(n=115) | No SRCC<br>(n=596) | Univariate<br>p-value | Multivariate†<br>OR (95% CI) | p-value |
| --- | --- | --- | --- | --- | --- |
| **Clinical** |  |  |  |  |  |
| Age at diagnosis, median (IQR) | 46.1 (32.3-60.7) | 47.8 (32.7-57.6) | 0.885 |  |  |
| Male carriers (%) | 48 (41.7) | 226 (37.9) | 0.425 |  |  |
| IGCLC criteria met* (%) | 35/87 (40.2) | 166/389 (42.7) | 0.745 |  |  |
| NCCN criteria met* (%) | 76/87 (87.4) | 315/389 (81.0) | 0.674 |  |  |
| Smoking ever* (%) | 36/113 (31.8) | 121/574 (21.1) | 0.236 |  |  |
| Heavy alcohol consumption* (%) | 5/111 (4.5) | 19/566 (3.4) | 0.408 |  |  |
| PPI use, ever* (%) | 32/102 (31.3) | 159/531 (29.9) | 0.613 |  |  |
| **Endoscopic** |  |  |  |  |  |
| # Total biopsies taken at endoscopy, median (IQR) | 38 (30-55) | 30 (20.5-45) | 0.066 | 1.01 (1.00-1.01) | 0.024 |
| Endoscopic findings (%) |  |  |  |  |  |
| Thickened mucosal folds | 7 (6.1) | 7 (1.2) | 0.002 | 3.57 (1.09-12.50) | 0.036 |
| Pale spots | 24 (20.8) | 66 (11.1) | 0.041 |  |  |
| Erythema | 27 (23.4) | 87 (14.6) | 0.049 |  |  |
| Friability | 2 (1.7) | 5 (0.8) | 0.239 |  |  |
| Nodularity | 19 (16.5) | 34 (5.7) | 0.001 | 2.56 (1.23-5.26) | 0.011 |
| Polyp(s) | 28 (24.3) | 114 (19.1) | 0.449 |  |  |
| Erosions | 5 (4.3) | 29 (4.8) | 0.775 |  |  |
| Ulcers | 3 (2.6) | 18 (3.0) | 0.953 |  |  |
| Mass | 3 (2.6) | 2 (0.3) | 0.011 | 5.88 (1.02-33.33) | 0.047 |
| **Histologic** |  |  |  |  |  |
| Intestinal Metaplasia (%) | 15 (13.1) | 38 (6.3) | 0.003 | 2.63 (1.45-4.76) | 0.001 |
| Gastritis (%) | 27 (23.4) | 230 (38.6) | 0.001 | 0.44 (0.28-0.69) | <0.001 |
| H. pylori (%) | 0 | 40 (6.7) | N/A† |  |  |
| Fundic gland polyp(s) (%) | 17 (14.7) | 59 (9.9) | 0.251 |  |  |

SRCC – signet ring cell carcinoma; IGCLC - International Gastric Cancer Linkage Consortium; IQR – interquartile range; NCCN – National Comprehensive Cancer Network; PPI – proton pump inhibitors
\*data missing for some carriers
†model could not estimate effect due to complete separation
‡model included the following variables: thickened mucosal folds, nodularity, mass, gastric intestinal metaplasia, gastritis, number of biopsies taken

### Carriers who had surveillance exams

Overall, 150 (40.2%) carriers underwent surveillance EGDs (≥2 EGDs) with a median follow-up of 28.6 months (IQR 15.1-56.3). **Supplementary Table 2** compares the characteristic of these carriers to those who only had baseline EGD. The surveillance cohort was older at diagnosis (median age 48.7 vs. 41.3, p=0.020) and had a lower proportion of patients undergoing gastrectomy (34% vs. 68.4%, p<0.001). There were no significant differences in endoscopic findings between the groups, and while both groups had similar overall detection rates of SRCCs (26%-29%), those who continued surveillance were less likely to have had SRCCs on their index EGD (4.7% vs. 29.8%, p<0.001).

### Surgical data

A total of 196 carriers (52.4%) underwent TG. Of these, 36 (18.3%) had no SRCC foci, 137 (69.9%) had stage T1a, and 11 (5.6%) had stage >T1a SRCC on the surgical specimen (12 had missing data on depth of invasion). Advanced lesions included stage T1bN0 (n=1), T1aN1 (n=1), T2N0 (n=1), T3N0 (n=4) and T4N0 (n=4). Ten (90.9%) of these carriers had visible lesions reported on endoscopy including erythema, nodularity, friability, thickened mucosal folds, pale mucosal spots with abnormal vascular pattern, erosions and/or ulcers and a visible mass lesion. The one individual without endoscopic findings was 8 years status post gastric bypass surgery and no visible lesions were reported in the gastric pouch. None of the carriers in the surveillance cohort who eventually underwent TG were found to have advanced-stage cancers, whereas all 11 advanced cases occurred in the cohort who had only baseline exams (8.5% of those undergoing surgery in this group).

Comparison of clinical, endoscopic and histologic features between cases with advanced versus localized stages are shown in **Table 3**. Endoscopically, thickened mucosal folds, erythema, nodularity, and gastric masses were more frequently observed in the group with advanced stage disease on univariate analysis. Histologically, identification of SRCC foci on the first baseline EGD and higher number of foci on endoscopic biopsies were factors significantly associated with advanced disease on univariate analysis. Average number of foci on EGDs in the advanced group was 2.5 foci compared to 0.67 foci in the localized group (**Supplementary Figure 1A**). On multivariate analysis, gastric masses or SRCC foci found on first EGD were associated with the advanced stage, though confidence intervals were wide reflecting small numbers.

**Table 3.**
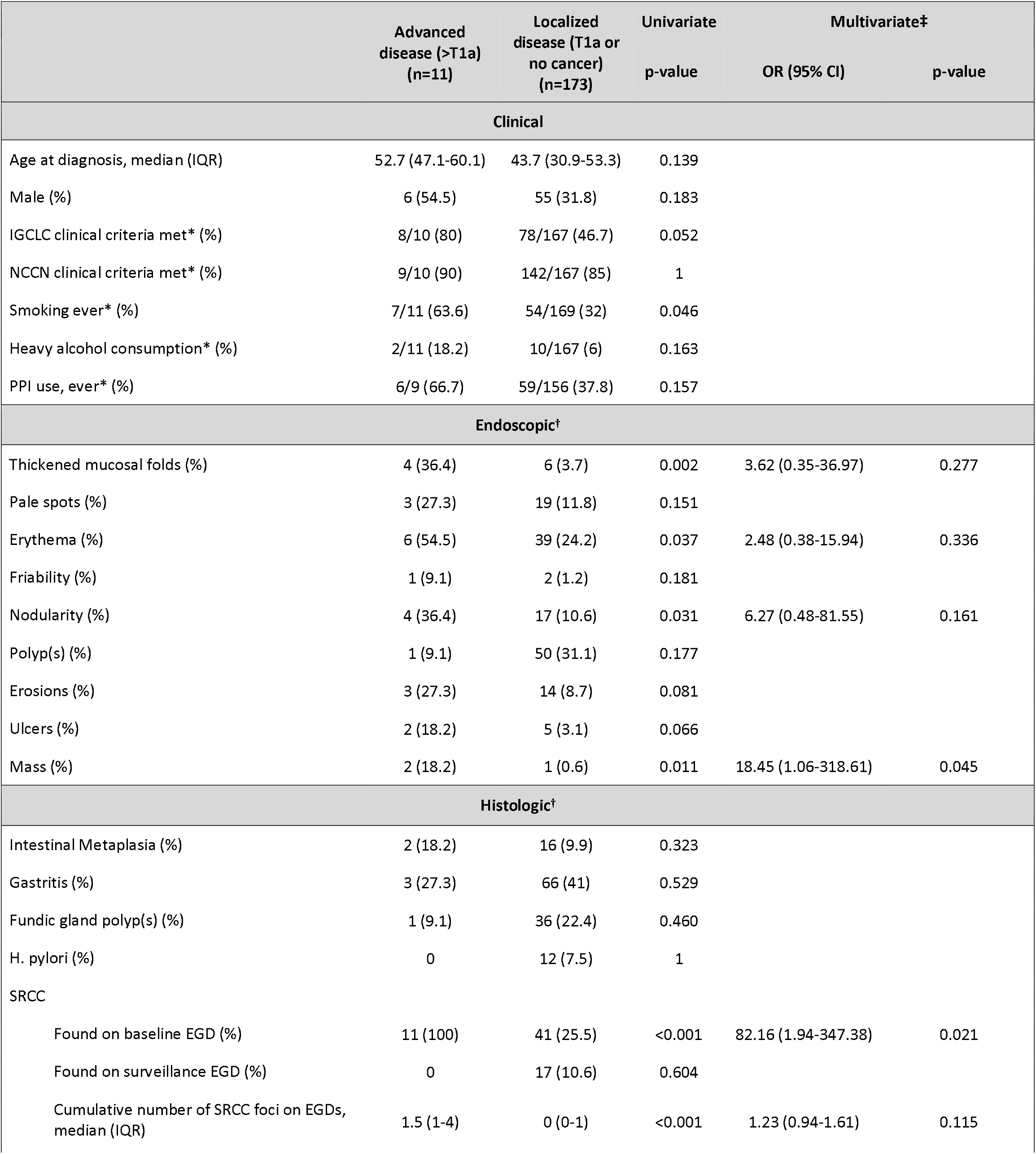
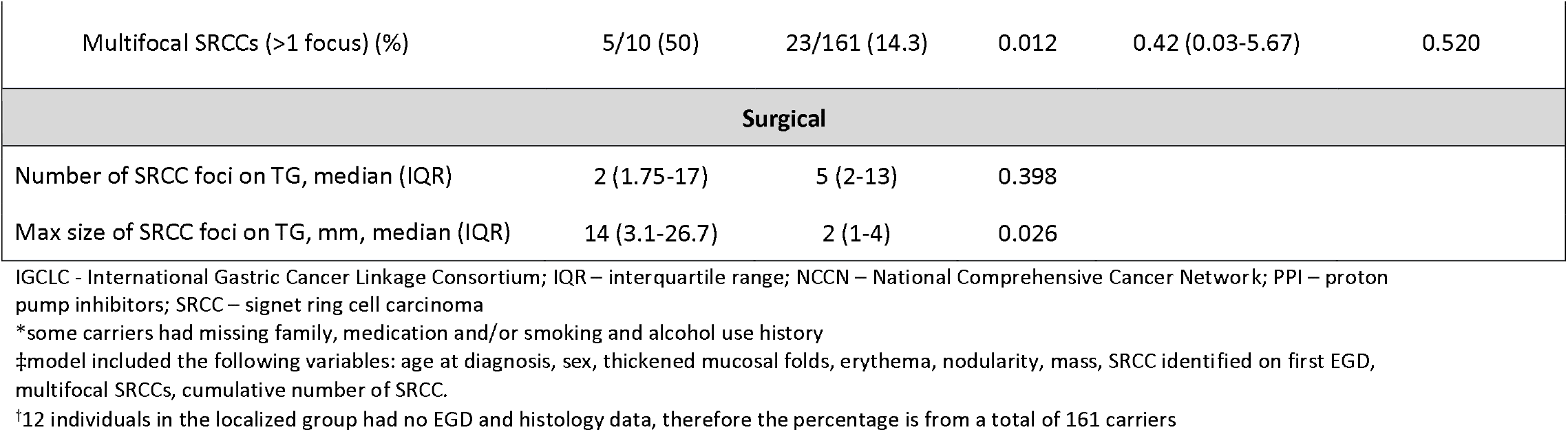
Comparison of clinical, endoscopic, histologic and surgical factors in *CDH1* carriers with vs. without advanced disease (>T1a) on gastrectomy specimen.

Figure 2 illustrates the sensitivity and specificity, as well as positive and negative predictive values (PPV/NPV) of the features associated with advanced disease. Sensitivity and specificity varied across features. Identification of SRCC on baseline endoscopy was the most sensitive feature (0.81) but had the lowest specificity (0.74), whereas masses and thickended folds demonstrated the highest specificities (0.99 and 0.96, respectively) but low sensitivity (0.18 and 0.36). Other features including erythema, nodular mucosa and multifocal SRCC demonstrated modest sensitivity (0.27-0.54) and intermediate specificity (0.75-0.89). NPVs were consistently high across all features (0.94-1.0), indicating strong ability to rule out advanced disease when these findings are absent. In contrast, PPVs were generally low (0.13-0.21), with the exception of masses (0.66) and, to a lesser extent, thickened folds (0.40), highlighting the limited ability of most features to reliably rule in advanced disease.

A median of 5 SRCC foci were found on TG specimens with a median size of 2mm. The foci in the advanced group were larger in size than those in the localized group (median of 14mm vs. 2mm, p=0.026). Both the number and size of these foci correlated weakly with the number of foci identified on EGD biopsies (Spearman rho 0.22 for number and 0.19 for size, p=0.017 and 0.048, respectively. **Supplementary Figure 1B, 1C**).

**Figure 2.**
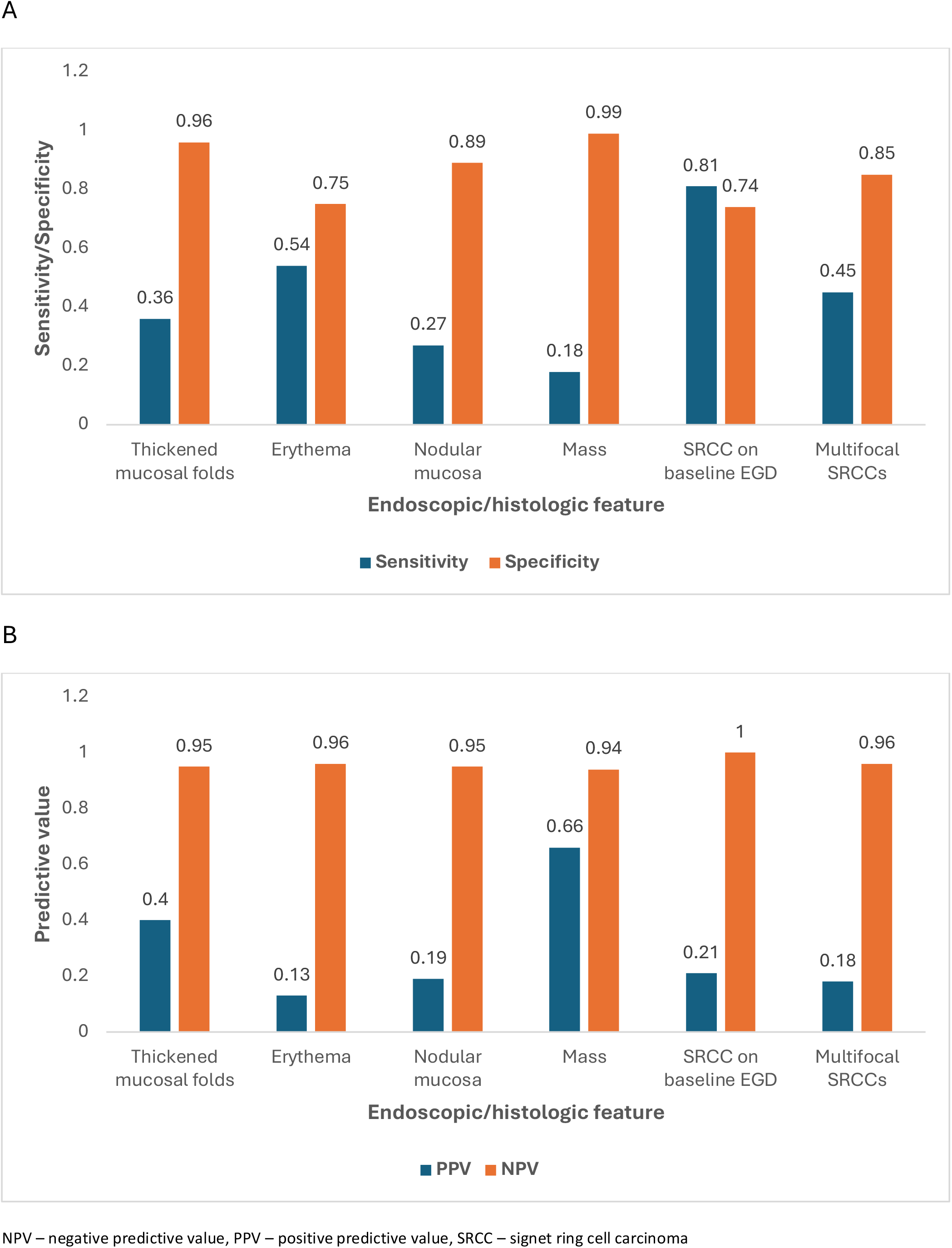
Diagnostic performance of endoscopic and histologic features for advanced disease. **A.** Sensitivity and specificity of endoscopic and histologic features. **B**. Positive predictive value (PPV) and negative predictive value (NPV) of the same features. All evaluated features had high NPVs (0.94–1.0), making them reliable for ruling out advanced disease. PPVs were modest (highest: masses 0.66, thickened folds 0.40), so they are less useful for ruling in disease. SRCC detection on baseline endoscopy was the most sensitive (0.81) but less specific (0.74), indicating it detects most cases but cannot definitively confirm disease.

### Oncologic data

Eleven carriers were found to have advanced stage cancer on their TG specimen (**Table 4)**. Seven carriers were asymptomatic prior to diagnosis of gastric cancer, two had epigastric pain and three had signs or symptoms of gastrointestinal bleeding. Four received neoadjuvant chemotherapy, 7 received adjuvant therapy, and 4 did not receive any therapy beyond surgery - one of whom developed recurrence 2 years after surgery and was then treated with chemotherapy. Six carriers have no evidence of disease (NED) after a median of 4.5 years from surgery (range 0.5-10 years). Four carriers, initially diagnosed without metastatic disease or lymph node involvement, developed recurrence at a median of 1.5 years after surgery, all 4 succumbed to their malignancy after a median of 3.7 years from diagnosis. One carrier was NED 8 months after surgery but passed away 3 months later from unknown cause.

**Table 4.** Oncologic outcomes of carriers with advanced (>T1a) disease.

| Carrier | Age (range) and sex | Pre surgery signs/symptoms | Endoscopic findings | Stage | Therapy | Outcome |
| --- | --- | --- | --- | --- | --- | --- |
| 1 | 60s, Male | Melena | Thickened folds, erythema, pale spots | T4aN0M0 | Adjuvant chemoradiation | NED 8 months after surgery. Died 3 months after last follow – no records available for cause of death. |
| 2 | 60s, Male | Asymptomatic | Erythematous friable mucosa, erosions, ulcer | T1bN0M0 | None | NED 2.5 years after surgery |
| 3 | 20s, Female | Epigastric pain, hematemesis | Mass | T4aN0M0 | None | NED 10 years after surgery |
| 4 | 50s, Female | Asymptomatic | Thickened folds, erythema | T3N0M0 | Neoadjuvant and adjuvant chemo | Recurrence 3 years after surgery. Died 5.5 years after diagnosis. |
| 5 | 60s, Female | Asymptomatic | None | T1aN1M0 | Neoadjuvant and adjuvant chemo | NED 3 years after surgery |
| 6 | 50s, Male | Asymptomatic | Thickened folds, erythematous friable mucosa | T2N0M0 | Adjuvant chemo | NED 4.5 years after surgery |
| 7 | 40s, Male | Asymptomatic | Erythema, pale spots | T3N0M0 | None | NED 7 years after surgery |
| 8 | 30s, Male | Epigastric pain | Nodular mucosa | T3N0M0 | Neoadjuvant chemo, adjuvant chemoradiation | Recurrence 1 year after surgery. Died 4 years after diagnosis |
| 9 | 40s, Female | Anemia (hemoglobin 6.6) | Thickened folds, ulcer, mass | T4aN0M0 | Neoadjuvant and adjuvant chemo | Developed malignant ascites while on chemo. Died 3.4 years after diagnosis |
| 10 | 50s, Female | Asymptomatic | Erosions | T3N0M0 | None after surgery. Chemo after recurrence | Recurrence 2 years after surgery. Died 3 years after diagnosis |
| 11 | 30s, Male | Asymptomatic | Nodular mucosa, pale spots with abnormal vascular pattern | T4aN0M0 | Adjuvant chemoradiation | NED 6 months after surgery |

NED – no evidence of disease

## Discussion

Management of *CDH1* carriers is challenging because predicting who will develop advanced gastric cancer and would benefit from prophylactic TG versus those who can continue endoscopic surveillance is not well established. Here, we present the first report from the GASTRIC consortium on clinicopathological features associated with localized and advanced SRCC in a large cohort of *CDH1* carriers from North American academic centers.

Among *CDH1* carriers with SRCC foci detected on endoscopy, 60% were identified on the baseline exam with most found to be T1a stage, present on random biopsies and located in the proximal stomach, in line with previous studies^7,8,12^. Several endoscopic features were significant independently associated with presence of SRCC foci on gastric biopsies including thickened mucosal folds, nodularity and gastric masses. Thickened folds can be a feature of linitis plastica, an infiltrative manifestation of diffuse gastric cancer^13^, while nodularity or masses are less commonly reported in diffuse gastric cancer. Erythematous mucosa, previously described in *CDH1* carriers with SRCC foci^7,14^, was significantly associated with SRCC detection on univariate analysis only. Similarly, mucosal pale spots which were previously reported as the most common mucosal lesion associated with SRCC foci^7,8^, were significant only on univariate analysis.

Histologically, GIM was found more commonly in endoscopies with SRCC foci, while gastritis showed an inverse association. One possible explanation is that GIM could mimic SRCCs, as the mucin containing goblet cells in GIM can resemble the mucin-filled cytoplasm of SRCCs^15^, while a significant inflammatory infiltrate could obscure SRCC making diagnosis more difficult. Alternatively, a connection between *CDH1*, GIM and gastric cancer is biologically plausible as *CDH1* promoter methylation and reduced E-cadherin expression have been reported in both GIM and early gastric cancers^16,17^. Additionally, an inverse association between immune cells and SRCCs may be supported by previous findings demonstrating that elevation of CD4+ lymphocytes in tumor microenvironment of *CDH1* carriers may limit the development of carcinogenesis and represent an immune response directed against the tumor^6^.

In the GASTRIC consortium, over 70% of carriers found to have SRCCs on EGD opted to undergo TG. Those who continued surveillance were less likely to have SRCC foci identified on their baseline EGD. This likely reflects older clinical guidelines, as our cohort included *CDH1* carriers with median year of diagnosis of 2018, when detection of SRCCs commonly prompted recommendation for total gastrectomy^1,18^, and is consistent with the Cambridge prospective cohort, where 5% of carriers without SRCC foci underwent surgery compared to 55% with positive biopsies^7^. None of the carriers under surveillance died from cancer, and none of those who eventually had prophylactic TG had advanced cancers, consistent with previous studies^7,8^, though the median follow-up of these carriers was only 24 months.

Nearly all *CDH1* carriers with advanced disease presented with endoscopic abnormalities, including thickened folds, pale spots, erythema, erosions, ulcers and masses. The only individual without overt endoscopic abnormalities was 8 years after gastric bypass surgery, possibly obscuring mucosal abnormalities due to post-surgical changes. The identification of SRCC foci on baseline, but not surveillance EGD and increased number of foci were the only significant pathologic findings associated with advanced disease. These findings may reflect a higher tumor burden, supported by the observed, albeit weak, correlation between the number and size of SRCC foci identified on TG specimens and the number of foci detected on EGD.

Collectively, these data support a management algorithm in which TG is recommended when mucosal abnormalities are identified and targeted biopsies of these lesions are positive for SRCCs^8^. Our study strengthens the evidence for this recommendation, with a substantially larger cohort of advanced cases (n=11) than previous reports^8,19^. Sensitivity and specificity varied across endoscopic and histologic features, highlighting differences in their ability to detect versus discriminate advanced disease. Identification of SRCC on baseline endoscopy was the most sensitive feature, whereas masses and thickened folds were the most specific but less sensitive, with other features demonstrating intermediate performance. Consistent with this, NPVs were uniformly high (0.94–1.0), indicating that absence of these findings is strongly reassuring. In contrast, PPVs were modest (0.13–0.66), limiting the ability of these features to reliably confirm advanced disease. These results suggest that patients with no identifiable endoscopic lesions may be safely managed with continued surveillance, whereas identification of mucosal lesions should prompt discussion of TG, especially in the setting of positive targeted biopsies. The role of multifocal SRCC foci on repeat endoscopies remains uncertain, as this parameter was non-significant on multivariate analysis. In the Cambridge cohort, multidocal foci were associated with higher number of SRCC foci on gastrectomy specimens, these these were all localized T1a lesions^19^.

Four carriers with advanced disease (40%) died of cancer complication. All four had visible lesions on their index EGD and two had signs and symptoms of gastrointestinal bleeding or epigastric pain, emphasizing the importance of evaluation for signs and symptoms of gastric cancer in *CDH1* carriers. Small retrospective studies did not identify advanced cases during follow up, nor had any cases of cancer deaths^20,21^. The large prospective studies from the NIH and Cambridge had only 3 cases with advanced cancer all of whom also had visible lesions on EGD and underwent therapeutic TG, with favorable postoperative outcomes, although follow-up was limited^8,22^. Additionally, in a recent observational multicenter study from Spain, the only patient diagnosed with advanced disease was found through targeted biopsy from a suspicious lesion detected on their baseline EGD^12^.

This study has several strengths. To our knowledge, this is the largest cohort to report on the outcomes of *CDH1* carriers across multiple academic centers. Importantly, our cohort includes 11 patients with advanced disease identified, substantially more than prior studies, which typically reported only one to two such cases^8,12,22^. This larger number of advanced cases enabled more meaningful comparisons between patients with advanced and localized disease. In addition, this is the first study to apply GEE models to address repeated endoscopic findings across multiple consecutive EGDs within the same individual, accounting for within-patient correlation. A number of limitations are acknowledged. Retrospective data did not allow for uniform reporting of endoscopic or pathological features and central review of pathology specimens was not feasible within the current study model. To mitigate variability, specific endoscopic and histologic features were prespecified prior to data collection to ensure consistency across sites. In addition, a relatively small number of carriers had longer-term endoscopic surveillance limiting evaluation of surveillance outcomes. Despite being a large cohort, numbers of advanced cancers were low which limits power to assess for significant associations.

In conclusion, in this large multi-center cohort, a minority of *CDH1* carriers developed advanced cancer, none of which were found during surveillance. Mucosal abnormalities were identified in nearly all cases advanced disease. Absence of endoscopic or histologic abnormalities was strongly reassuring, whereas the presence of abnormalities had modest positive predictive value, and their significance for ruling in advanced disease remains uncertain. Long-term follow up of *CDH1* carriers undergoing endoscopic surveillance is required to validate predictors of advanced disease. Additionally, future studies should explore the role of molecular markers^23^, the immune microenvironment^6^ and artificial intelligence tools^24^ to better stratify risk *CDH1* carriers, ultimately guiding decisions on which individuals require surgery and those patients who could safely be managed with endoscopic surveillance.

## Authors contribution

OG, BD, SDR, AS, MJH, XL, AB, SGP, REB, MER, PPS, ES, BWK, MA, SSK: conceptualization, methodology. OG, GG: analysis.

OG: software.

OG, CMD, EK, CS, AG, SB, MH, TA, MJ, KG, BD, EK, CL, MS, SS, DC, BAL, GH, YC, AK, SE, KH: data curation, resources.

OG, SSK: writing – original draft.

OG, CMD, EK, GG, CS, AG, SB, MH, TA, MJ, KG, BD, EK, CL, MS, SS, DC, BAL, GH, YC, AK, SE, KH, SDR, AS, MJH, XL, AB, SGP, REB, MER, PPS, ES, BWK, MA, SSK: writing – review and editing.

SSK: supervision

## Ethics statement

All procedures followed were in accordance with the ethical standards of the responsible committee on human experimentation (institutional and national) and with the Helsinki Declaration of 1964 and later versions.

## Role of funding source

This research received no external funding.

## Declaration of interests

All authors declare no competing interests.

## Data sharing

Data from this study can be requested from OG. Data access requires an approved proposal detailing intended use and a signed data-sharing agreement. Data will be available from publication, and no time limit applies to data availability.

## Abbreviations

EGD: esophagogastroduodenoscopy
GEE: Generalized estimating equation
IGCLC: International Gastric Cancer Linkage Consortium
IQR: interquartile range)
NCCN: National Comprehensive Cancer Network)
NED: no evidence of disease
PPI: proton pump inhibitor
SD: standard deviation
SRCC: signet ring cell carcinoma
TG: total gastrectomy

## Notes

### Competing Interest Statement

The authors have declared no competing interest.

### Author Declarations

The study was approved by the institutional review boards of all centers: University of Chicago, Sinai Health System, University of Pennsylvania, University of Michigan, The Ohio State University, University of Pittsburgh, University of Colorado, University of Kansas, Yale University, Fox Chase Cancer Center, University of California San Francisco and Columbia University.

