## Supplementary Table 1; Supplementary Table 2; Supplementary Figure 1 for "Clinicopathological Factors Associated with Gastric Signet Ring Cell Carcinoma in *CDH1* Pathogenic Variant Carriers: Report from the GASTRIC Consortium"

**Supplementary Table 1**: 2020 IGCLC testing criteria and the 2025 NCCN guidelines

| 2020 IGCLC Guidelines: |
|---|
| Family criteria:<br><br>• ≥2 cases of gastric cancer in family regardless of age, with at least one diffuse gastric cancer (DGC)<br>• ≥1 case of DGC at any age and ≥1 case of lobular breast cancer (LBC) <70 years in different family members<br>• ≥2 cases of LBC in family members <50 years<br><br>Individual criteria:<br><br>• DGC <50 years<br>• DGC at any age in individuals of Māori ethnicity<br>• DGC at any age in individuals with a personal or family history (1st degree) of cleft lip/cleft palate<br>• History of DGC and LBC, both diagnosed <70 years<br>• Bilateral LBC, diagnosed <70 years<br>• Gastric *in situ* signet ring cells and/or pagetoid spread of signet ring cells in individuals <50 years |
| 2025 NCCN Guidelines: |
| • Individual with a known *CDH1* pathogenic variant in the family<br>• An individual with DGC at any age<br>• Family history of ≥2 first-degree or second-degree relatives with gastric cancer with at least one diagnosed at age ≤50 years or at least one confirmed to be DGC at any age<br>• Individual meeting criteria for *CDH1* testing based on NCCN Guidelines for high-penetrance breast cancer susceptibility Genes |

**Supplementary Table 2:** Comparison of *CDH1* carriers with screening EGD only (1 EGD) vs. surveillance EGDs (≥2 EGDs)

| | Screening EGD (n=188) | Surveillance EGDs (n=150) | p-value |
|---|---|---|---|
| Age at *CDH1* diagnosis, median (IQR) | 41.3 (29.2-54.6) | 48.7 (33.3-57.4) | 0.020 |
| Male sex (%) | 75 (39.9) | 51 (34) | 0.266 |
| Surveillance, months, median (IQR) | 5.7 (3.3-11.3) | 28.6 (15.1-56.3) | <0.001 |
| Testing criteria met (%) | | | |
| IGCLC | 72 (38.3) | 58 (38.6) | 0.825 |
| NCCN | 147 (78.2) | 123 (82) | 0.237 |
| Number of EGDs, mean ±SD | 1 | 3.6 ±1.9 | <0.001 |
| Carriers with EGDs with mucosal abnormalities (%) | 9 (4.8) | 4 (2.7) | 0.314 |
| Thickened mucosal folds | 41 (21.9) | 44 (29.3) | 0.113 |
| Erythema | 16 (8.5) | 20 (13.3) | 0.153 |
| Nodularity | 5 (2.7) | 10 (6.7) | 0.076 |
| Ulcer | 4 (2.1) | 0 | 0.132 |
| Mass | | | |
| Carriers with positive SRCC (%) | | | |
| Overall | 56 (29.8) | 40 (26.7) | 0.527 |
| On first screening EGD only | 56 (29.8) | 7 (4.7) | <0.001 |
| On targeted biopsies | 13 (7) | 7 (4.7) | 0.378 |
| On random biopsies | 47 (25) | 36 (24) | 0.810 |
| Cumulative number of foci, median | 0 (0-4) | 0 | 0.509 |
| (IQR) | 0.43 ±0.9 | 0.56 ±1.7 | 0.509 |
| Cumulative number of foci, mean ±SD | | | |
| Carriers undergoing TG (%) | | | |
| Overall | 129 (68.6) | 51 (34) | <0.001 |
| Localized (stage 0 or T1a) | 112 (87.5) | 50 (98.1) | 0.035 |
| Advanced (stage>T1a) | 11 (8.9) | 0 | 0.035 |
| Age at TG, median (IQR) | 41.2 (31.6-54.1) | 49.9 (37.7-57.8) | 0.059 |

IGCLC - International Gastric Cancer Linkage Consortium; IQR – interquartile range; NCCN – National Comprehensive Cancer Network; SD – standard deviation; SRCC – signet ring cell carcinoma; TG – total gastrectomy

**Supplementary Figure 1:** Correlation between number of SRCC foci identified on EGD and A. advanced stage disease (p<0.001); B. number of foci on gastrectomy specimen (rho 0.22, p=0.017); C. maximal size of foci on gastrectomy (rho 0.19, p=0.048)

A

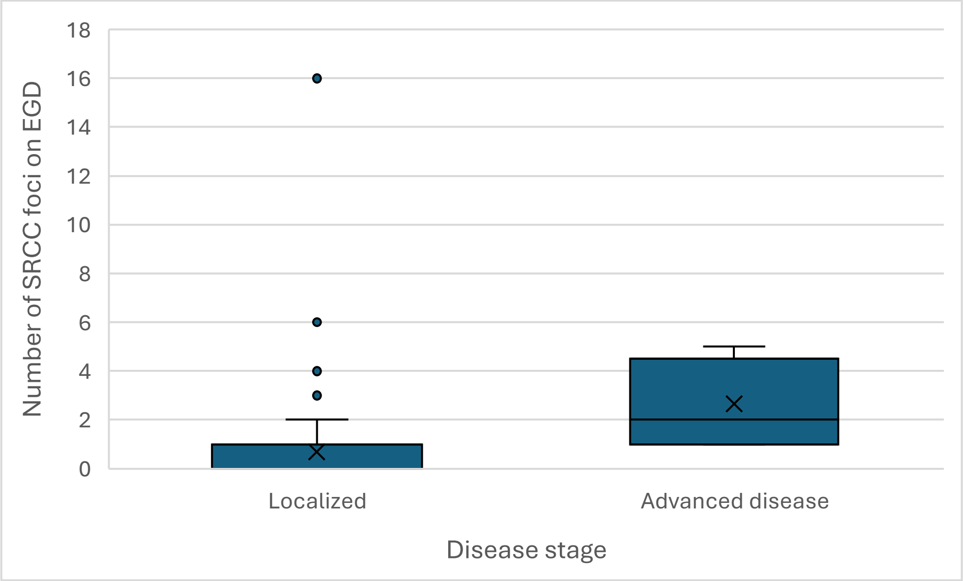


B

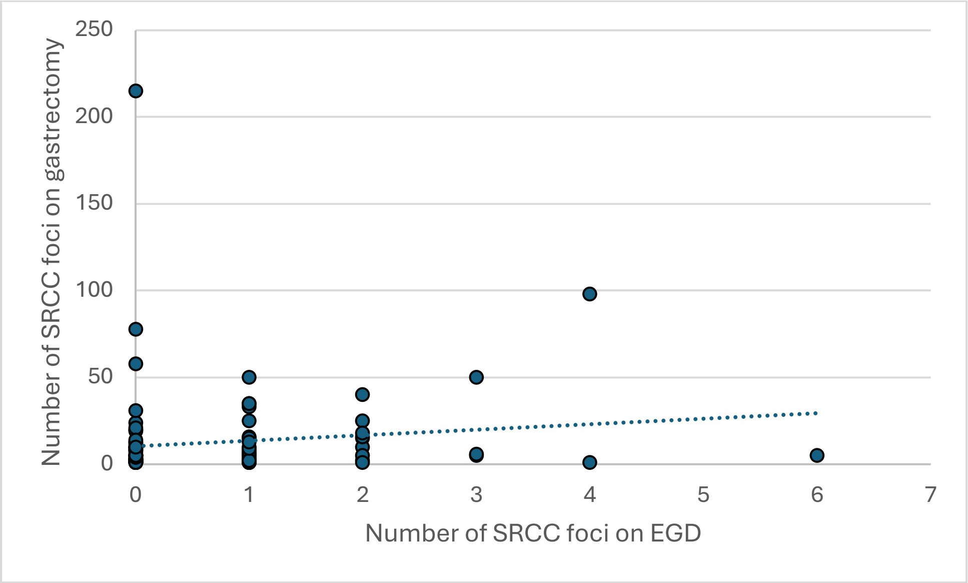

C

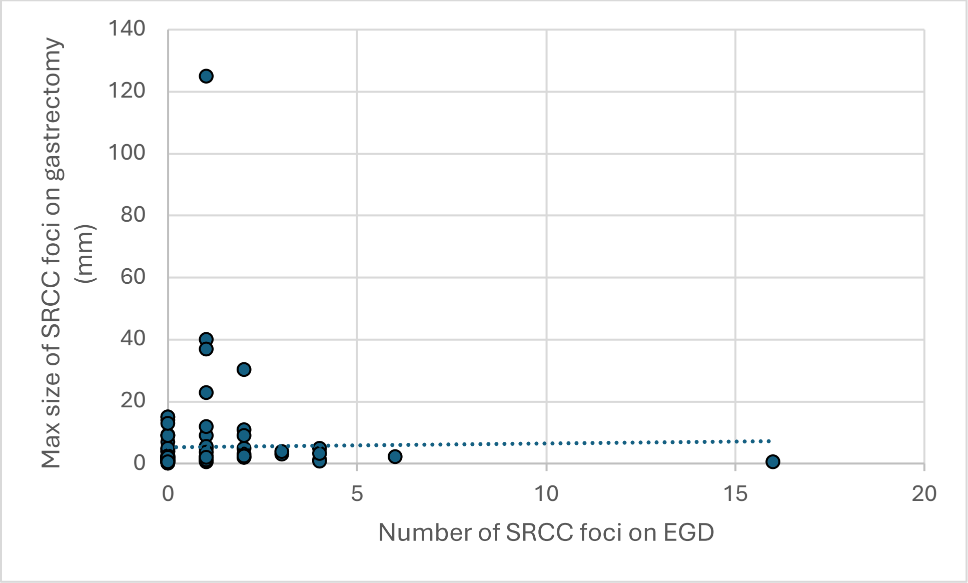

Max size of SRCC foci on gastrectomy (mm)
140
120
100
80
60
40
20
0
0
5
10
15
20
Number of SRCC foci on EGD
